# Repetitive transcranial magnetic stimulation to the motor cortex leads to a sequential increase in phase synchronization and power of TMS-evoked electroencephalographic recordings

**DOI:** 10.1101/2024.04.24.24306176

**Authors:** Enrico De Martino, Adenauer Girardi Casali, Bruno Andry Nascimento Couto, Thomas Graven-Nielsen, Daniel Ciampi de Andrade

## Abstract

**Background:** High-frequency (10 Hz) repetitive transcranial magnetic stimulation (rTMS) to the primary motor cortex (M1) is used to treat several neuropsychiatric disorders, but its main mechanism of action remains unclear.

**Objective:** To probe four cortical hubs used for rTMS (M1; dorsolateral-prefrontal cortex, DLPFC; anterior cingulate cortex, ACC; posterosuperior insula, PSI) with TMS coupled with high-density electroencephalography (TMS-EEG) and measure cortical excitability and oscillatory dynamics before and after active and sham rTMS to M1.

**Methods:** Before and immediately after active or sham M1-rTMS (15 min, 3,000 pulses at 10 Hz), single-pulse TMS evoked EEG were recorded at the four targets in 20 healthy individuals. Measures of cortical excitability and oscillatory dynamics were extracted at the main frequency bands (α [8-13 Hz], low-β [14-24 Hz], high-β [25-35 Hz]).

**Results:** Comparing active and sham M1 rTMS, M1 TMS-EEG demonstrated an increase in high-β synchronization in electrodes around M1 stimulation area and remotely in the contralateral hemisphere (p=0.026). The increase in high-β synchronization (48-83 ms after TMS-EEG stimulation) was succeeded by an enhancement in low-β power (86-144 ms after TMS-EEG stimulation) both locally and in the contralateral hemisphere (p=0.006). No significant differences were observed in TMS-EEG responses probing DLPFC, ACC, or PSI.

**Conclusion:** M1-rTMS engaged a sequence of enhanced phase synchronization, followed by an increase in power occurring within M1, that spread to remote areas and was measurable after the end of the stimulation session. These results are relevant to understanding the M1 neuroplastic effects of rTMS and associated changes in cortical activity dynamics.

## INTRODUCTION

High-frequency (10 Hz) repetitive transcranial magnetic stimulation (rTMS) to the primary motor cortex (M1) is a non-invasive neuromodulation technique able to induce analgesic effects [1] and has therapeutic potentials in chronic pain, stroke rehabilitation, and movement disorders, among others [2]. Although still unclear, rTMS-induced analgesia may provoke long-lasting cortical plastic changes by repetitively depolarizing myelinated axons in M1 [3], probably via Hebbian synaptic plasticity mechanisms [4]. Previous studies based on motor-evoked potentials (MEPs) and intra- cortical excitability demonstrated that M1 rTMS increased corticomotor excitability [5] and normalized reduced intra-cortical excitability in chronic pain [6]. Importantly, M1 is highly connected to cognitive and somatosensory networks, including interoceptive and nociceptive top-down modulatory areas [7], and it is thus currently assumed that M1 stimulation has significant modulatory effects in extra-motor corticospinal networks [8].

Recent advancements in TMS-compatible electroencephalography (TMS-EEG) have opened the possibility of directly assessing cortical responses to a probing pulse of TMS both at the stimulation site and remotely, which can open new perspectives in studying the rTMS mechanisms. TMS-EEG allows for the measurement of cortical excitability and connectivity with enough temporal resolution to early and later evoked responses in motor and extra-motor areas [9]. The technique involves the application of sub-threshold TMS single pulses to a targeted cortical area under the recording of EEG to assess the ensuing changes in cortical neural activity [10]. Averaged cortical responses, known as TMS-evoked EEG potentials (TEPs), are waveforms derived from time-locked and phase-locked EEG segments to TMS pulses, which are particularly effective for examining cortical excitability [11]. Furthermore, TMS-EEG also provides the opportunity to explore the frequency content of evoked cortical oscillations both locally in the stimulated area and globally across cortical regions connected to the stimulated cortical target [9].

In M1, the dominant oscillation is within the β-band [12,13], which involves pyramidal neurons, as evidenced by corticomuscular coherence [14]. However, M1 also expresses an important oscillatory activity within the α-band, also termed mu-rhythm, which is related to the integration of somatosensory stimuli in a manner like the modulation of visual perception by occipital α oscillations [15]. Previous studies combining TMS-EEG to M1 with continuous theta burst stimulation (TBS) have shown a significant increase in power in beta frequency [16] and a reduction in alpha power and phase synchronization in the stimulation site [16,17]. By contrast, intermittent TBS has reported a decrease in power in α-band frequencies in the electrodes located away from the stimulation site [18].

Furthermore, studies combining TMS to M1 with functional resonance magnetic imaging (fMRI) have also demonstrated that TMS led to BOLD changes in functionally connected cortical non-motor regions, such as the insular, prefrontal, and cingulate regions [19,20]. However, the fine-grained temporal dynamics of these correlations and their directionality remain largely unknown.

Here, we investigated whether M1-rTMS influenced cortical excitability and oscillatory dynamics within the α- and β-bands in healthy individuals by probing motor and extra-motor (dorsolateral prefrontal, anterior cingulate, and posterior insular) cortices with TMS-EEG in a sham- controlled setting.

## METHODS

### Participants

This study included 20 healthy adults (12 females). Age, height, and weight (mean ± SD) were 25±4 years, 173±12.6 cm, and 67±15 kg. None of the participants were on medications, and the exclusion criteria were non-systemic diseases and neuropsychiatric disorders, known pregnancy, and any contraindications to TMS [21]. The local ethics committee approved the study (N-20220018), and the protocol was registered at ClinicalTrials.gov (NCT05714020). The sample size was determined based on the calculation of the difference between two dependent means. Based on the analgesic effect of rTMS applied to M1 (a reduction of 1.5 points in pain intensity on the numeric rating scale with a standard deviation of 1.8 at the end of the treatment [22]), an effect size of active rTMS to M1 was estimated to be 0.83. Using G*Power for statistical power analysis with a power of 0.80, alpha level of 0.05, and effect size of 0.80, a minimum of 14 participants was necessary.

### Study design

The present study involved two experimental sessions separated by one week. In the first visit, participants were randomly assigned to either sham or active rTMS to the left M1, with 10 participants receiving sham rTMS first. All participants received the other rTMS protocol during the second visit. Both active and sham rTMS procedures, as well as participant instructions, were kept consistent across groups. Before and after the rTMS intervention, three TMS-EEG assessments on three distinct left cortical areas were performed: First, the dorsolateral prefrontal cortex (DLPFC) and M1 regions were stimulated in a randomized order in all 20 participants (10 receiving DLPFC stimulation first). In order to collect the post-measurement assessments within 1 hour after rTMS, half of the participants underwent TMS-EEG to the anterior cingulate cortex (ACC), while the second half underwent the posterosuperior insula (PSI). Each TMS-EEG protocol took approximately 8 minutes, and 5-minute breaks were ensured between runs. MEPs were assessed both before and after TMS-EEG measurements.

### Repetitive Transcranial Magnetic Stimulation

Magstim Super Rapid2 Plus1 stimulator (Magstim Company Ltd) with a figure-of-eight-shaped coil (70-mm Double Air Film Coil) was used for rTMS (15 min of stimulation, targeting the hot spot of the right first dorsal interosseous (FDI) muscle, 30 trains of 10-s pulses at 10 Hz frequency and 20-s intervals between trains, totaling 3000 pulses) [2]. Stimulation intensity was 90% of the resting motor threshold (rMT). For sham stimulations, a coil identical in size, color, shape, and mimicking the active coil sound (70-mm double air film sham coil) was used.

### Corticospinal excitability

Silver chloride electrodes (Ambu Neuroline 720) were placed on the right FDI muscle fibers. Single- pulse TMS was delivered using the rTMS device and a figure-of-eight shaped coil D70². The hotspot of the FDI muscle was determined as the coil position that evoked a maximal peak-to-peak MEP for a given stimulation intensity. The rMT was the lowest TMS intensity that could produce MEPs exceeding 50 μV in half of the trials [23]. Ten pulses were delivered at 120% and 140% of rMT.

### Electroencephalographic recordings of TMS-evoked potentials

Electroencephalograms were recorded using a TMS-compatible amplifier (g.HIamp EEG amplifier, g.tec medical engineering GmbH) with a passive electrode cap (64 electrodes, Easycap) placed according to the 10-5 system, with the Cz electrode on the vertex. The ground electrode was placed on the right zygoma, the online reference was on the right mastoid process, and two electrodes on the lateral side of the eyes recorded the electrooculogram. Electrode impedance was kept under 5 kΩ. Raw signals were amplified and sampled at a rate of 4800 Hz.

TMS was delivered using the same biphasic stimulator as used for rTMS with a figure-eight coil to stimulate DLPFC and M1 (D70² coil) and a double-cone coil (D110 cone-coil) to stimulate the ACC and PSI targets. During recordings, participants sat on an ergonomic armchair and were instructed to gaze at a fixation spot on the wall to reduce oculomotor muscle activity. The TMS-click sound masking toolbox (TAAC; [24]) with noise-cancelation in-ear headphones (ER3C Etymotic 50 Ohm) were used to mitigate auditory responses to TMS coil clicks. An EEG net cap (GVB-geliMED GmbH) with a plastic stretch wrap film was applied over the EEG cap to reduce somatosensory artifacts triggered by coil contact with electrodes.

TMS-neuronavigation (Brainsight TMS Neuronavigation, Rogue Research Inc.) was used to target the cortical spots between assessments. For M1, TMS-evoked potentials were obtained from motor hotspots (Fig. 1A) at 90% of rMT. The DLPFC target was identified on the middle frontal gyrus, based on the method described by Mylius et al. [25], with TMS intensity set to 110% of the rMT of the FDI muscle (Fig. 2A). The ACC target was identified 4 cm in front of the hotspot of the tibialis anterior (TA) muscle scalp representation [26] (Fig. 3A). The hotspot of the TA muscle was determined as the coil position that evoked a TA-evoked response for a given stimulation intensity.

**Figure 1:**
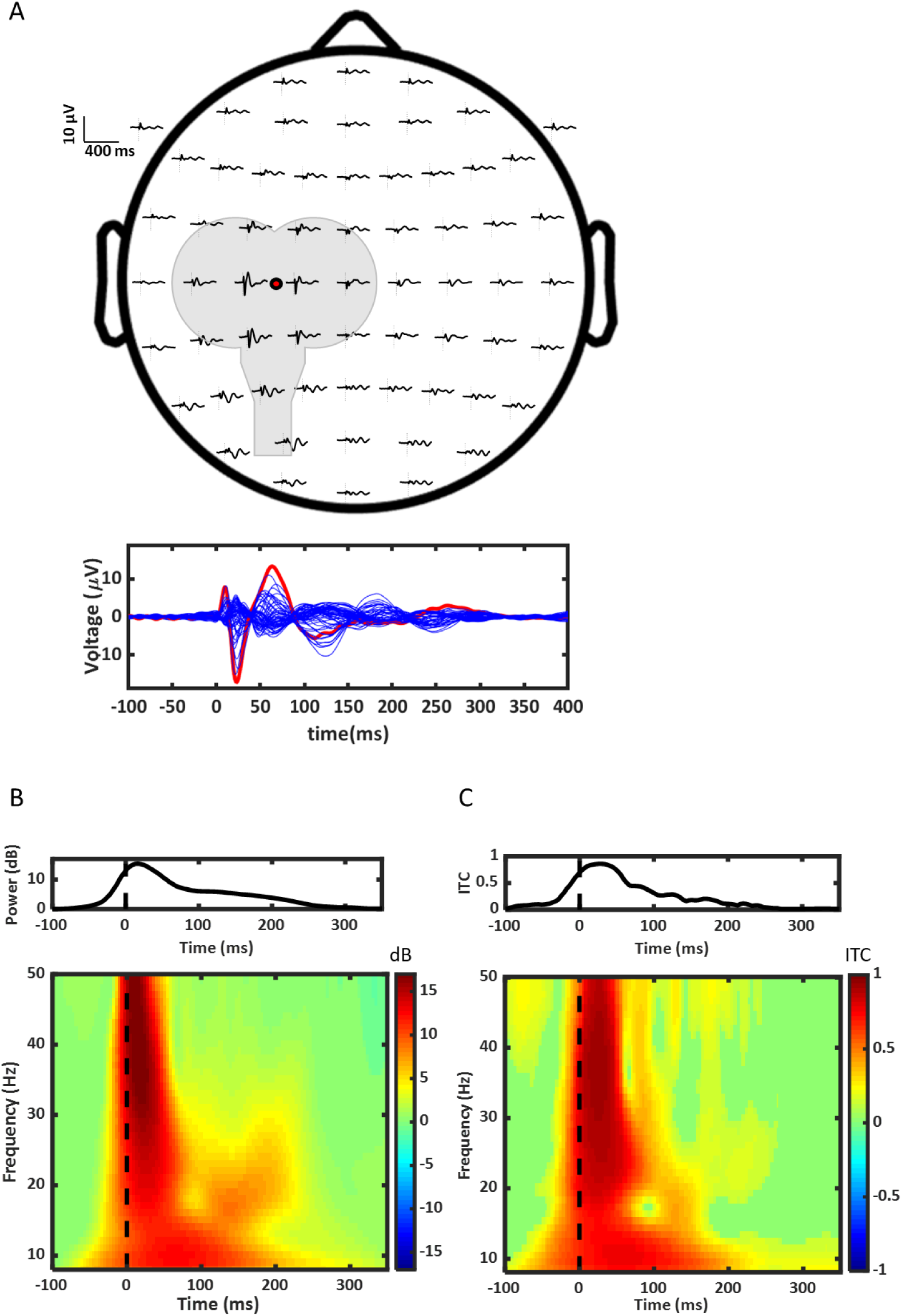
Sample data of transcranial magnetic stimulation (TMS)-evoked potentials recorded with EEG following single pulse stimulation to the left primary motor cortex (M1) in a representative participant. **A)** Topographical representation of M1 TEPs for each individual electrode. The red dot corresponds to the area of stimulation. The butterfly plot shown below depicts the superposition of TEPs for all electrodes. The red line corresponds to the C3 electrode, and the blue lines correspond to the other 62 channels. **B)** Mean broadband event-related spectral perturbation (ERSP) over time on C3 (time: from -100 to 350 ms). Below is the ERSP map calculated on the same electrode. **C)** Mean broadband intertrial coherence (ITC) over time on C3 (time: from -100 to 350 ms). Below is the ITC map calculated on the same electrode.

**Figure 2:**
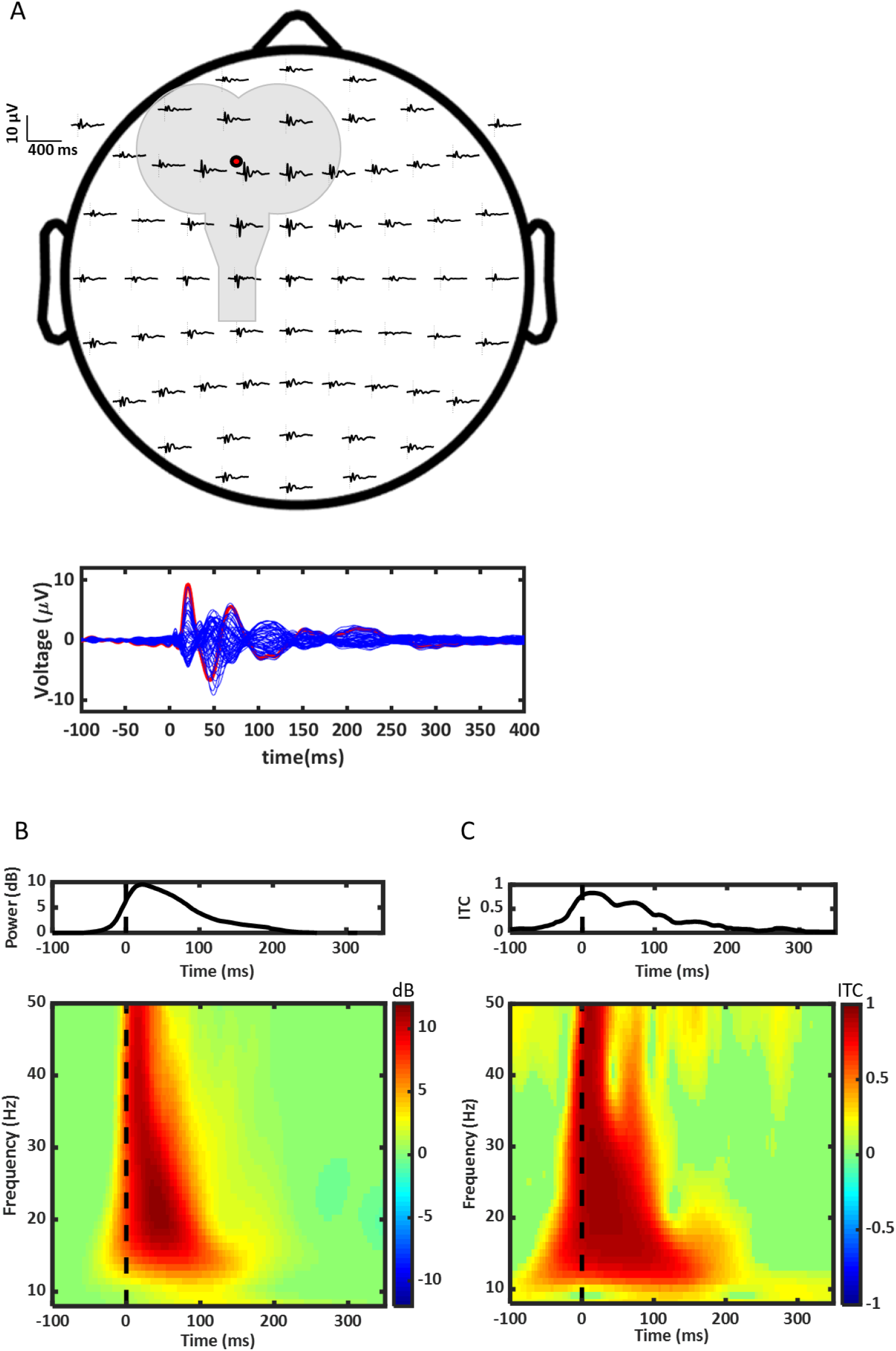
Sample data of transcranial magnetic stimulation (TMS)-evoked potentials recorded with EEG following single pulse stimulation to the left dorsolateral prefrontal cortex (DLPFC) in a representative participant. **A)** Topographical representation of DLPFC TEPs for each individual electrode. The red dot corresponds to the area of stimulation. The butterfly plot is shown below. The red line corresponds to the F1 electrode, and the blue lines correspond to the other 62 channels. **B)** Mean broadband event-related spectral perturbation (ERSP) over time on F1 (time: from -100 to 350 ms). Below is the ERSP map calculated on the same electrode. **C)** Mean broadband intertrial coherence (ITC) over time on F1 (time: from -100 to 350 ms). Below is the ITC map calculated on the same electrode.

**Figure 3:**
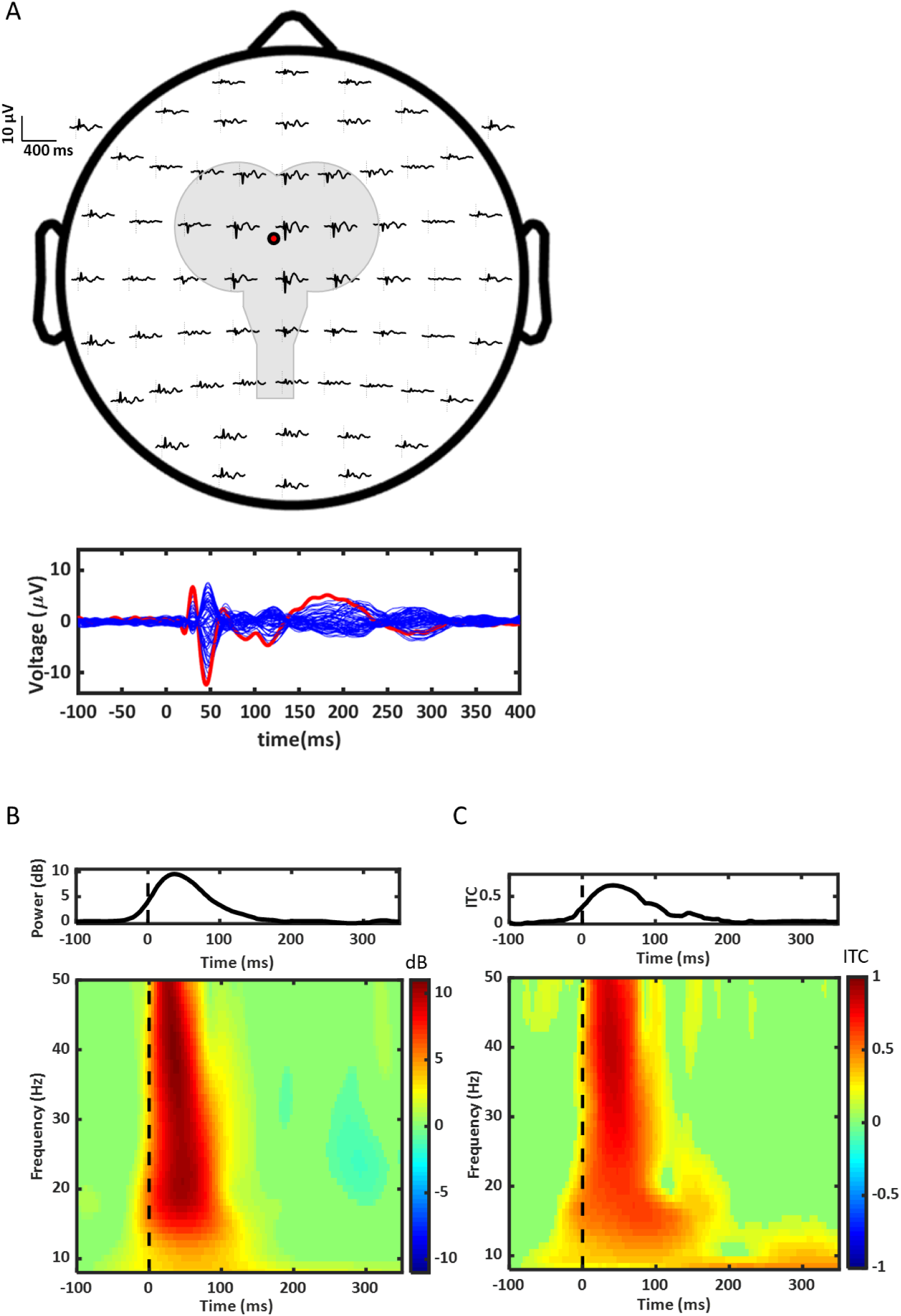
Sample data of transcranial magnetic stimulation (TMS)-evoked potentials recorded with EEG following single pulse stimulation to the anterior cingulate cortex (ACC) in a representative participant. **A)** Topographical representation of ACC TEPs for each individual electrode. The butterfly plot is shown below. The red line corresponds to the FCz electrode, and the blue lines correspond to the other 62 channels. **B)** Mean broadband event-related spectral perturbation (ERSP) over time on FCz (time: from -100 to 350 ms). Below is the ERSP map calculated on the same electrode. **C)** Mean broadband intertrial coherence (ITC) over time on FCz (time: from -100 to 350 ms). Below is the ITC map calculated on the same electrode.

The rMT of the TA muscle was determined as the lowest TMS intensity to produce visible muscle responses, and TMS-EEG was performed at 90% of the TA rMT. The PSI target was identified as previously described [27] (Fig 4A), and stimulation was similarly set at 90% of the TA rMT.

**Figure 4.**
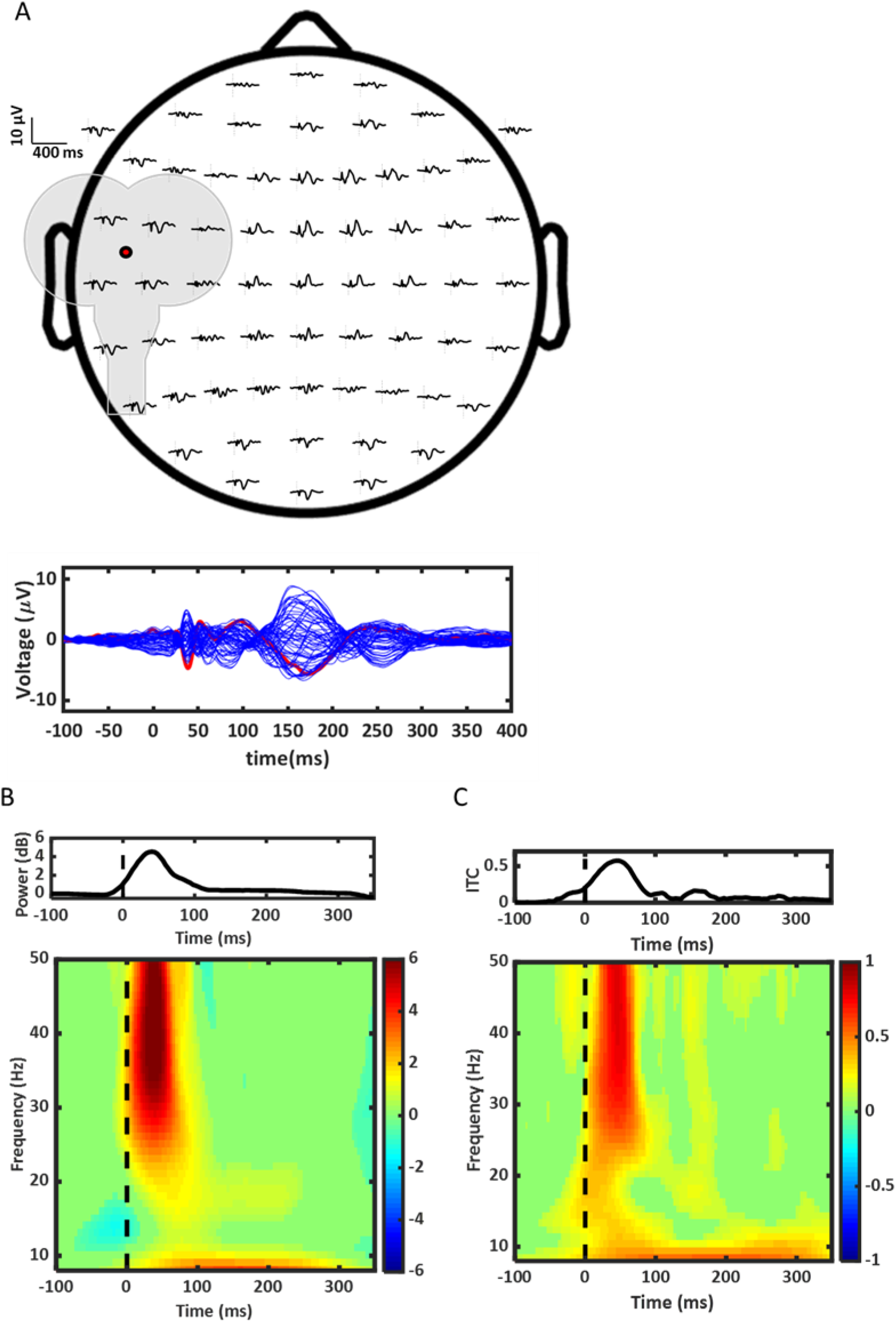
Sample data of transcranial magnetic stimulation (TMS)-evoked potentials recorded with EEG following single pulse stimulation to the posterosuperior insula (PSI) in a representative participant. **A)** Topographical representation of PSI TEPs for each individual electrode. The butterfly plot is shown below. The red line corresponds to the FC7 electrode, and the blue lines correspond to the other 62 channels. **B)** Mean broadband event-related spectral perturbation (ERSP) over time on FC7 (time: from -100 to 350 ms). Below is the ERSP map calculated on the same electrode. **C)** Mean broadband intertrial coherence (ITC) over time on FC7 (time: from -100 to 350 ms). Below is the ITC map calculated on the same electrode.

A real-time visualization tool (rt-TEP) was used to ensure detectable TMS-evoked potentials in all cortical targets [10]. This allowed for monitoring the quality of the recordings and allowed for minor adjustments in TMS coil angulation and orientation, ensuring the presence of early peak-to- peak TMS-evoked potentials (average of 20 trials) at the nearest electrode to the stimulation area. The TMS-neuronavigation and rt-TEP were utilized throughout the study to monitor the TMS coil location and the highest signal-to-noise ratio in EEG recordings. Approximately 160-180 pulses were administered for each condition, with interstimulus intervals randomly jittered between 2600 and 3400 ms [28].

Pre-processing was performed using customized algorithms based on the EEGlab toolbox [29] running on Matlab R2019b (The MathWorks). EEG signals were segmented into trials of 1600 ms around the TMS pulse, which occurred at time zero (±800 ms). In the M1 TMS-EEG epoch, a segment of the pre-TMS EEG signal (-11 to -3 ms) was used to substitute the peri-TMS EEG recordings from -2 to 6 ms [30]. The same procedure was applied for deep TMS targets (ACC and PSI) in a larger peri-TMS interval (0-20 ms) to adapt to the double-cone coil electric field. Epochs and channels with noise, eye blinks, eye movements, or muscle artifacts were identified and removed. The EEG data were band-pass filtered (1-80 Hz, Butterworth, 3rd order), down sampled to 1200 Hz, re-referenced to average reference, baseline corrected, and merged for the two conditions (Pre- and Post-rTMS). Independent component analysis (ICA, EEGLAB runica function) was applied to the combined dataset to remove additional residual artifacts [13]. The dataset was divided into the original Pre- and Post-rTMS conditions, and the epochs were re-segmented to the window of ±600 ms surrounding the TMS pulse. Lastly, signals from any disconnected or high-impedance channels were interpolated using spherical splines [29].

To assess the global cortical excitability, global-mean field power (GMFP) was calculated as the root-mean-squared value of the TEP across all electrodes in the 20-300 ms time interval after TMS stimulation [13]. To assess the local cortical excitability, the local mean field power (LMFP) was calculated across the electrodes close to the TMS coil in the 20-300 ms time interval after TMS stimulation [13]. For M1 stimulation, C1, C3, Cp3, Cp1 electrodes were selected, likewise for DLPFC (AF3, F3, F1, FC3, FC1), ACC (FCz, Cz, FC1, FC2, C1, C2), and PSI (FC7, F C3, C7, C5, C3).

Time-frequency maps were extracted between 8 and 45 Hz using Morlet wavelets with 3.5 cycles, as implemented in the EEGLAB toolbox and previously reported [31]. The following TMS- evoked EEG parameters were extracted in the time-frequency domain:

- Event-related spectral perturbation (ERSP) was calculated to quantify power amplitudes in the frequency domain. ERSP was computed from the time-frequency maps as the average spectral power ratio of individual EEG trials relative to the pre-stimulus period (-600 to -50 ms). ERSP allows the identification of the changes in power as a function of time and frequency [9,31]. The significance of ERSP maps with respect to the baseline was assessed by bootstrapping samples from the pre-stimulus period (500 permutations, two-sided comparison, p-value < 0.05 after false discovery rate (FDR) correction for multiple comparisons). Mean power spectra were then calculated by averaging significant ERSP values across electrodes and time samples (Fig. 1B, 2B, 3B, 4B).
- Inter-trial coherence (ITC) was extracted as a measure of phase synchronization. ITC was calculated by normalizing the complex-valued single-trial time-frequency values by their corresponding moduli and taking the absolute value of the across-trials averaged results. The significance of ITC maps with respect to the baseline was assessed by bootstrapping samples from the pre-stimulus period (500 permutations, one-sided p-value < 0.05 after FDR), and significant ITC values were averaged across electrodes, time samples, and frequency bands (Fig. 1C, 2C, 3C, 4C).

To investigate if the observed changes in ERSP and ITC were not due to volume conduction from a common source activity, the weighted Phase Lag Index (wPLI) was also calculated. wPLI assesses the asymmetry of the phase difference distribution between pairs of EEG signals, which is indicative of phase synchronization between electrodes free from zero-lag components. For each session, wPLI was calculated as described by Vinck et al. (2011) [32]. The resulting connectivity matrix was then averaged across each time window of interest for both electrodes belonging to the same cluster (intra- cluster) and for different clusters (inter-cluster). The clusters and time windows were chosen based on a cluster analysis (details in the *Statistical Analysis* section).

### Statistical analysis

Matlab and Statistical Package for Social Sciences (SPSS, version 25; IBM) were used for statistical analyses. Data are presented as mean ± standard deviation. For cortical excitability (global and local mean filed power), phase-based connectivity analysis (wLPI), and MEPs amplitude, Student’s paired t-tests were used to compare the absolute changes from pre-stimulation (Post- vs. Pre-rTMS) between active and sham stimulations. For the time-frequency analysis, three distinct frequency bands, α (8– 13 Hz), low-β (14–24 Hz), and high-β (25-35 Hz), were selected a priori, and a spatiotemporal group- level comparison of ERSP and ITC, averaged across frequency bands, was performed between active and sham rTMS using a non-parametric permutation test, corrected for multiple comparisons through cluster-based statistics [33] as implemented in the open-source FieldTrip Toolbox [34]. The test consisted of two levels: first-level t-statistics and cluster-level statistics. The first-level statistics quantifies the effect at each spatiotemporal sample, establishing a threshold for identifying samples as members of clusters. Spatiotemporal samples with first-level statistics considered significant (p<0.05; two-sided) were grouped based on temporal and spatial adjacency (minimum of two channels per cluster). The sum of first-level statistics within each cluster was used as cluster-level statistics and compared to the maximum distribution of values obtained after randomizing data across types of stimulations (Monte Carlo approximation with 5000 random permutations). The cluster analysis was applied to subject-normalized ERSP and ITC maps, which were constructed by subtracting the pre-rTMS (active or sham) individual maps from the corresponding post-rTMS maps. Clusters were significant when the observed summed statistics exceeded 95% of the values resulting from random permutations.

## RESULTS

All participants completed all TMS-EEG assessments and both sham and active rTMS to M1 without adverse effects. Repetitive TMS intensities were 62.1±8.3% for sham and 62.5±8.1% for active rTMS. The TMS-evoked potential intensities for each cortical area and the average number of artifact-free epochs are reported in Supplementary Tables 1 and 2. Data from three subjects were excluded due to TMS-EPs peak-to-peak amplitude not reaching 6 μV [10].

### Effects of M1 rTMS on M1 TMS-EEG and MEPs

Local and global mean field power analyses did not show significant differences between active and sham rTMS (Supplementary Fig. 1). Data-driven analyses for group-level comparisons of ERSP and ITC revealed a difference in the high-β band in the ITC in the time interval 48-83 ms. Furthermore, in the low-β band, ERSP also revealed a difference in the time interval 86-144 ms. Topographic plots for M1 TMS probing revealed the electrodes where differences were present (Fig. 5), allowing their grouping into distinct clusters and two equal time intervals: early (48-83 ms) and late (86-144 ms) intervals. A significant increase in high-β band early (48-83 ms) phase reset (ITC) after active rTMS was detected compared to sham in the left central cluster (t(16) = 3.258; p = 0.005; electrodes: C3, C5, Cp3, and Cp5), in the left frontal cluster (t(16) = 2.446; p = 0.026; electrodes: Af3, F1 and F3), and in the right frontal cluster (t(16) = 4.052; p = 0.001; electrodes: Af4, F2 and F4). These changes were temporally followed (86-144 ms) by an increase in lower-β band later power (ERSP) after active rTMS compared with sham in the left centro-parietal cluster (t(16) = 2.943; p = 0.009; electrodes: C1, C3, Cp1, Cp3, P1, and P3), right centro-parietal cluster (t(16) = 3.683; p = 0.002; electrodes: C2, C4, Cp2, Cp4, P2, and P4), and left prefrontal cluster (t(16) = 4.684; p = 0.001; electrodes: Fp1, Af3, and Af7) (non-normalized parameters are reported in Table 1 and 2).

**Figure 5:**
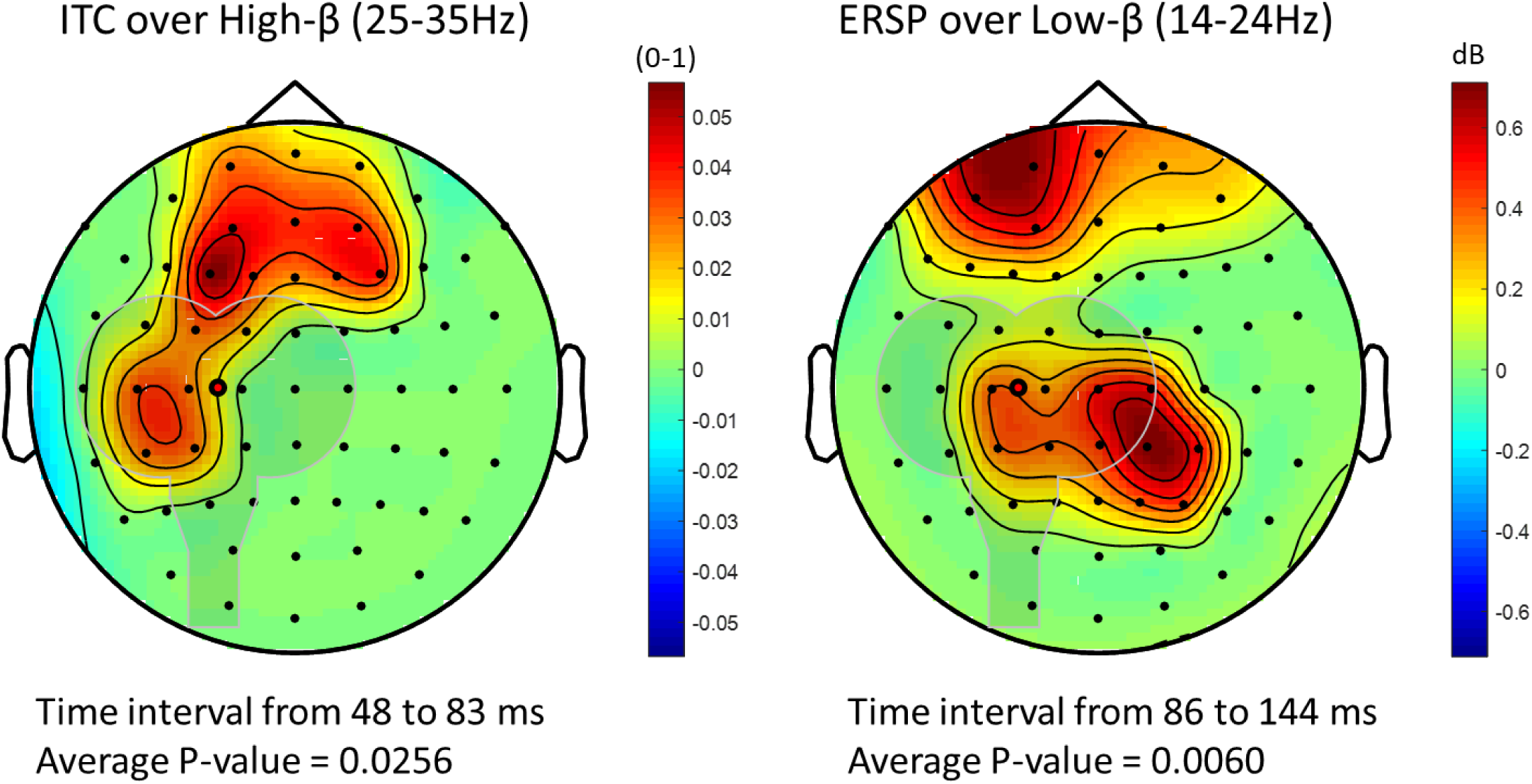
Topographic maps of the average difference between active and sham rTMS for the spatiotemporal clusters of ITC (left, high-β band) and ERSP (right, low-β band) found significant. Average P-values and time intervals of the significant clusters are displayed below.

**Table 1.** Mean ± standard deviation of the high-β band inter-trial coherence (0-1) for each condition before and immediately after repetitive transcranial magnetic stimulation (rTMS) to the primary motor cortex. Two regions in the 48-83 ms time interval were selected based on significant effects when analyzing the differences in non-parametric permutation tests.

| Cluster | Condition | Pre-rTMS | Post-rTMS | Absolute change |
| --- | --- | --- | --- | --- |
| Left central | Sham | 0.46±0.17 | 0.44±0.20 | -0.02±0.08 |
| | Active | 0.44±0.17 | 0.48±0.16 | 0.04±0.05 |
|  | p-value |  |  | 0.005 |
| Left frontal | Sham | 0.43±0.18 | 0.40±0.18 | -0.02±0.09 |
| | Active | 0.41±0.16 | 0.46±0.16 | 0.04±0.07 |
|  | p-value |  |  | 0.026 |
| Right frontal | Sham | 0.38±0.17 | 0.35±0.15 | -0.03±0.09 |
| | Active | 0.35±0.17 | 0.38±0.16 | 0.04±0.06 |
|  | p-value |  |  | 0.001 |

**Table 2.** Mean ± standard deviation of the low-β band event-related spectral perturbation (dB) for each condition before and immediately after repetitive transcranial magnetic stimulation (rTMS) to the primary motor cortex. Four regions in the 86-144 ms time interval were selected based on significant effects when analyzing the differences in non-parametric permutation tests.

| Cluster | Condition | Pre-rTMS | Post-rTMS | Absolute change |
| --- | --- | --- | --- | --- |
| Left centro-parietal | Sham | 1.76±1.15 | 1.27±1.06 | -0.49±0.69 |
| | Active | 1.74±1.50 | 1.75±1.19 | 0.01±0.69 |
|  | p-value |  |  | 0.009 |
| Left prefrontal | Sham | 1.17±1.01 | 0.83±0.81 | -0.35±0.50 |
| | Active | 1.07±0.82 | 1.33±0.66 | 0.26±0.56 |
|  | p-value |  |  | 0.001 |
| Right centro-parietal | Sham | 1.00±0.84 | 0.73±0.85 | -0.28±0.69 |
| | Active | 0.84±0.71 | 1.23±0.67 | 0.39±0.66 |
|  | p-value |  |  | 0.002 |

Phase-based connectivity analyses confirmed the sequential events described above in high-β band wPLI after active rTMS compared with sham between the left central cluster (peri-stimulation site) and left prefrontal cluster both at early (t(16) = 2.490; p = 0.024) and late (t(16) = 2.181; p = 0.044) time intervals. These findings were similarly followed by an increase in high-β (t(16) = 3.533; p = 0.003) and low-β (t(16) = 2.511; p = 0.023) band wPLI after active rTMS between the left central cluster (peri-stimulation site) and right centro-parietal cluster at the late time interval (non-normalized parameters are reported in Table 3). Absolute changes in MEP amplitudes were not significant (Supplementary Tables 3).

**Table 3.** Mean ± standard deviation of the weighted phase lag index (0-1) for each condition before and immediately after repetitive transcranial magnetic stimulation (rTMS) to the primary motor cortex. Two regions were selected based on significant effects when analyzing the differences in event- related spectral perturbation and inter-trial coherence.

| Clusters | Condition | Pre-rTMS | Post-rTMS | Absolute change |
| --- | --- | --- | --- | --- |
| High-β band: left central-left prefrontal (time interval 48-83 ms) | Sham | 0.35±0.13 | 0.32±0.10 | -0.03±0.08 |
| | Active | 0.32±0.12 | 0.35±0.13 | 0.03±0.07 |
|  | p-value |  |  | 0.024 |
| High-β band: left central-left prefrontal (time interval 86-144 ms) | Sham | 0.17± 0.05 | 0.14 ± 0.03 | -0.03±0.05 |
| | Active | 0.15 ± 0.06 | 0.17 ± 0.06 | 0.02±0.07 |
|  | p-value |  |  | 0.044 |
| Low-β band: left central-right centro-parietal (time interval 86-144 ms) | Sham | 0.21±0.07 | 0.18±0.05 | -0.03±0.07 |
| | Active | 0.18±0.04 | 0.21±0.05 | 0.02±0.05 |
|  | p-value |  |  | 0.023 |
| High-β band: left central-right centro-parietal (time interval 86-144 ms) | Sham | 0.15±0.05 | 0.14±0.03 | -0.01±0.04 |
| | Active | 0.14±0.03 | 0.17±0.04 | 0.03±0.04 |
|  | p-value |  |  | 0.003 |

### Effects of M1 rTMS on DLPFC, ACC, and PSI TMS-EEG

Local and global mean field power analysis (Supplementary Fig. 2, 3, and 4) and the time-frequency analyses did not show any significant difference between active and sham rTMS in any of the three other cortical areas proved with TMS-EEG.

## DISCUSSION

The present study provides original insights into both the local and remote connectivity changes immediately after a session of M1-rTMS at 10Hz. Initial increases in faster β-band intertrial coherence occurred in electrodes around M1 and in the ipsilateral frontal and homologous contralateral hemispheres. Initial increases in intertrial coherence were followed by increases in power in the slower β-band, observable locally in the prefrontal ipsilateral and peri-motor contralateral hemispheres. Phase-based connectivity analyses further supported that active rTMS increased phase lagging between the stimulated M1 area and remote extra-motor areas. Contrarily, cortical responses to extra-motor probing (DLPFC, ACC, and PSI) were not significantly affected by rTMS to M1, suggesting that connectivity changes were mainly measurable in M1-related networks.

### Effects of M1 rTMS on M1 β-band oscillatory activity

The current results demonstrated that active 10 Hz rTMS to M1 did not significantly enhance the α- band oscillation in electrodes close to the stimulation area via causal entrainment of brain oscillations similar to what has been observed in the parietal cortex [35] or in previous studies using continuous and intermittent TBS [16–18]. Instead, M1-rTMS increased high β-band oscillatory synchronization and low β-band oscillatory power. Previous studies combining TMS-EEG with continuous and intermitted TBS used TMS intensities above the rest of the motor threshold, which provokes muscle contractions. Consequently, the extent to which the reported changes were influenced by sensory reafference from the periphery, or the spinal cord is unclear. A previous study applying single pulses TMS or trains of rhythmic or arhythmic rTMS to M1 at ∼ 18 Hz (peak of individual participant’s resting-state beta oscillation) also triggered an increase in beta power on resting EEG, independently of the pattern of stimulation [36]. These results [36], and our own, corroborate the concept that beta oscillatory response after M1-rTMS reflects potentiation of the endogenous M1’s β-band natural oscillatory activity, regardless of rTMS frequency. This supports the idea that the after-effects of rTMS delivered at frequencies ∼ 10-20Hz are related to the M1 main frequency rather than to effects linked to the stimulation frequency band. This is in line with frequency ranges found to have therapeutic values for chronic pain management by M1 rTMS(see for review [1]).

An important finding of the current study is the increase in high β-band oscillatory synchronization after active rTMS. ITC is a measure computed from single-trial cortical responses, reflecting the temporal and spectral synchronization within the EEG response, and indicating the extent to which underlying phase-locking occurs, providing a direct measure of cortical synchrony [29]. High β-band oscillatory synchronization in cortical regions has previously been demonstrated to play a crucial role in interregional cortical communication and function, and their coordination across regions and inter-regional coordination jointly improve behavioral performance [37]. Thus, the increased high β-band oscillatory synchronization found in the current study could be hypothesized as an increase in communication-through-coherence between M1 and its connected areas [38]. The communication-through-coherence theory suggests that brain rhythms encompass distinctly increased excitation and inhibition phases, and inputs are most effective when timed to coincide with excitation phases and not with phases of inhibition [39]. This optimal timing can occur if the inputs are rhythmic, thereby influencing synchronized rhythms in the target brain regions [37]. Another main finding of the current study was the increase in high-β synchronization after active rTMS followed by an increase in low-β band power. This temporal interaction between cortical rhythms may indicate a cross-frequency coupling from a faster to a slower rhythmic state. It is well- known that cross-frequency coupling is a crucial mechanism for interaction between the many discrete frequencies of rhythm observable in neocortical networks [40]. In animal and human studies, phase–amplitude coupling has been observed, converging on the notion that it plays an important functional role in local computation and long-range communication in large-scale brain networks [41].

A final relevant finding of the current study was the increased β-band connectivity, as measured by wPLI, across several different clusters of electrodes after active rTMS. The phase-based connectivity analysis suggests that this effect was not produced by volume conduction and that the increase in β-band oscillatory synchronization and power after rTMS do not originate from the directly targeted cortex but also from remote cortical regions, allowing the inference of effective connectivity changes and driving the changes in its interconnected areas. This is supported by a large body of animal and human evidence [8,42] showing that pain analgesia and somatosensory effects of M1 stimulation are dependent on the engagement of extra-motor areas and diffuse effects such as the release of endogenous opioids [43]. In fact, it was suggested that M1 has areas that are highly connected to the extra motor (e.g., cognitive control, interoceptive, pain modulatory) network [44], which could be central to the clinical effects reported to date after M1 rTMS [22].

### Effects of M1 rTMS on DLPFC, ACC, and PSI oscillatory activity

No significant changes in cortical excitability or oscillations were found when TMS probed DLPFC, ACC, and PSI after M1 rTMS. Previous concurrent TMS-fMRI studies have shown that TMS can induce neurovascular responses in functionally connected non-motor areas, including the insula, cingulate cortex, frontal and parietal cortices [19,45]. A recent study has shown that TMS over M1, synchronized with fMRI acquisition, led to increased activation in the bilateral insula [20]. Additionally, dynamic causal modeling indicated direct inputs from M1 to the insula and ACC [46]. Despite this evidence of activation in connected cortical regions, our study did not capture any neuroplastic effects in these areas following 10 Hz rTMS to M1. This could be due to the transient nature of the effects or the need for multiple rTMS sessions to induce lasting neuroplastic changes.

### Limitations

The main limitation of the present study is the absence of behavioral assessment. This was an active choice in the design of the study, given that the connectivity changes after M1 rTMS were mainly unknown, while the behavioral effects after M1 stimulation have been previously described [47]. Furthermore, only M1 was targeted with rTMS. Considering that responsiveness to active rTMS may vary significantly across different cortical areas based on endogenous oscillation, future research should investigate the effects of targeting different cortical areas, such as DLPFC, ACC, or PSI, which could induce different cortical responses. Finally, only 10 participants underwent TMS-EEG targeting the ACC and PSI since the post-rTMS effect on M1 is short-lasting [48].

### Conclusions

Compared to sham-rTMS, active M1 rTMS engaged an enhanced TMS-synchronization, followed by an increase in TMS-evoked power amplitude occurring within M1 main frequencies and spreading to remote connected areas.

## CRediT author statement

**Enrico De Martino:** Conceptualization, Methodology, Formal analysis, Investigation, Data curation, Visualization, Writing- Original draft preparation; Writing - Review & Editing; Project administration; **Adenauer Girardi Casali**: Methodology, Software; Formal analysis; Data Curation; Visualization, Writing - Review & Editing; **Bruno Andry Nascimento Couto**: Software, Formal analysis, Data Curation, Visualization, Writing - Review & Editing, Visualization; **Thomas Graven- Nielsen**: Conceptualization, Methodology, Writing - Review & Editing, Funding acquisition; **Daniel Ciampi de Andrade**: Conceptualization, Methodology, Visualization, Writing- Original draft preparation; Writing - Review & Editing; Project administration, Funding acquisition.

## Data Availability

All data produced in the present study are available upon reasonable request to the authors

## SUPPLEMENTARY MATERIAL

**Supplementary figure 1.**
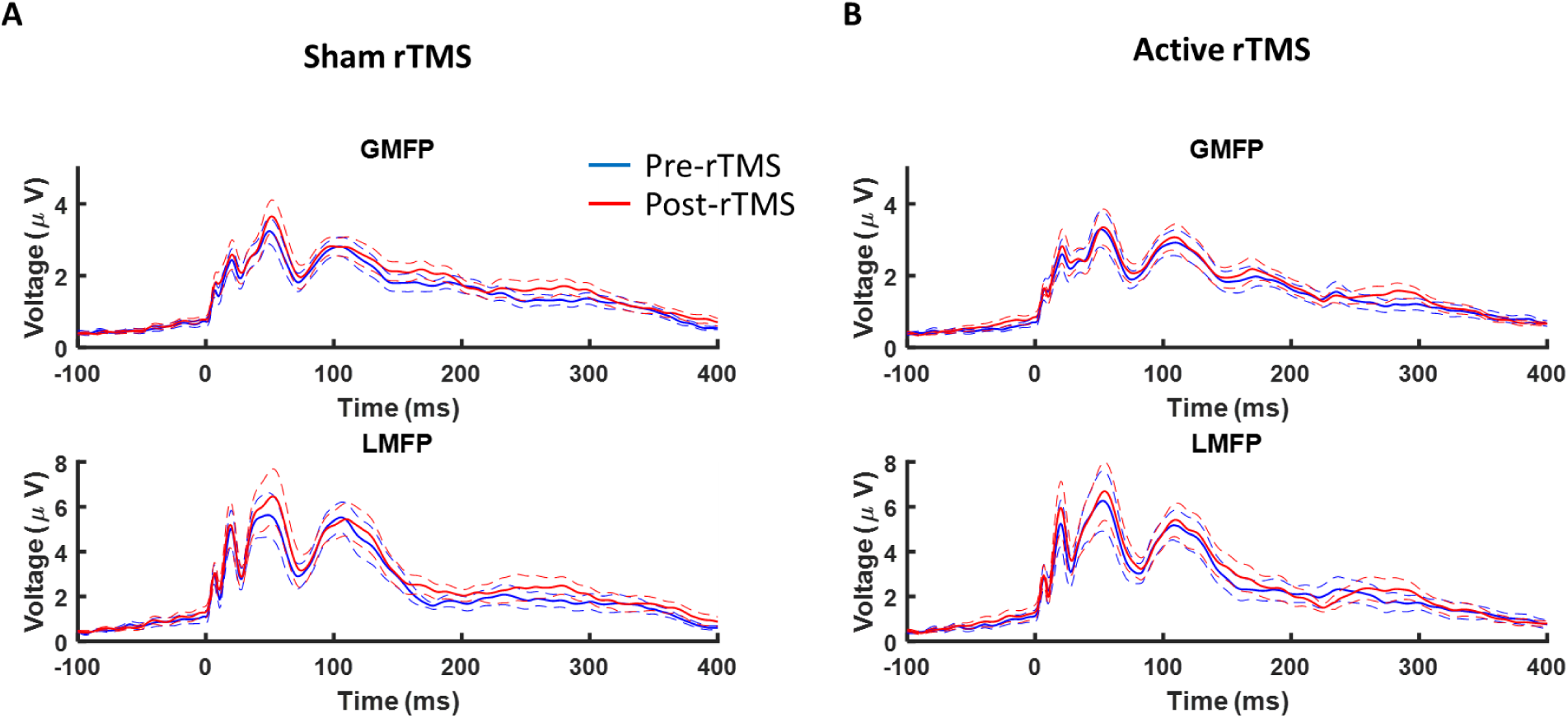
Global Mean Field Power (GMFP) and Local Mean Field Power (LMFP) from the primary motor cortex (M1) stimulation. Before repetitive transcranial magnetic stimulation (Pre-rTMS) is shown in blue, and immediately after repetitive transcranial magnetic stimulation (post-rTMS) is in red. Solid lines indicate the mean values, and dashed lines represent the standard deviation for each condition. **A)** Sham rTMS 10 Hz rTMS to M1; **B)** Active rTMS 10 Hz rTMS to M1.

**Supplementary figure 2.**
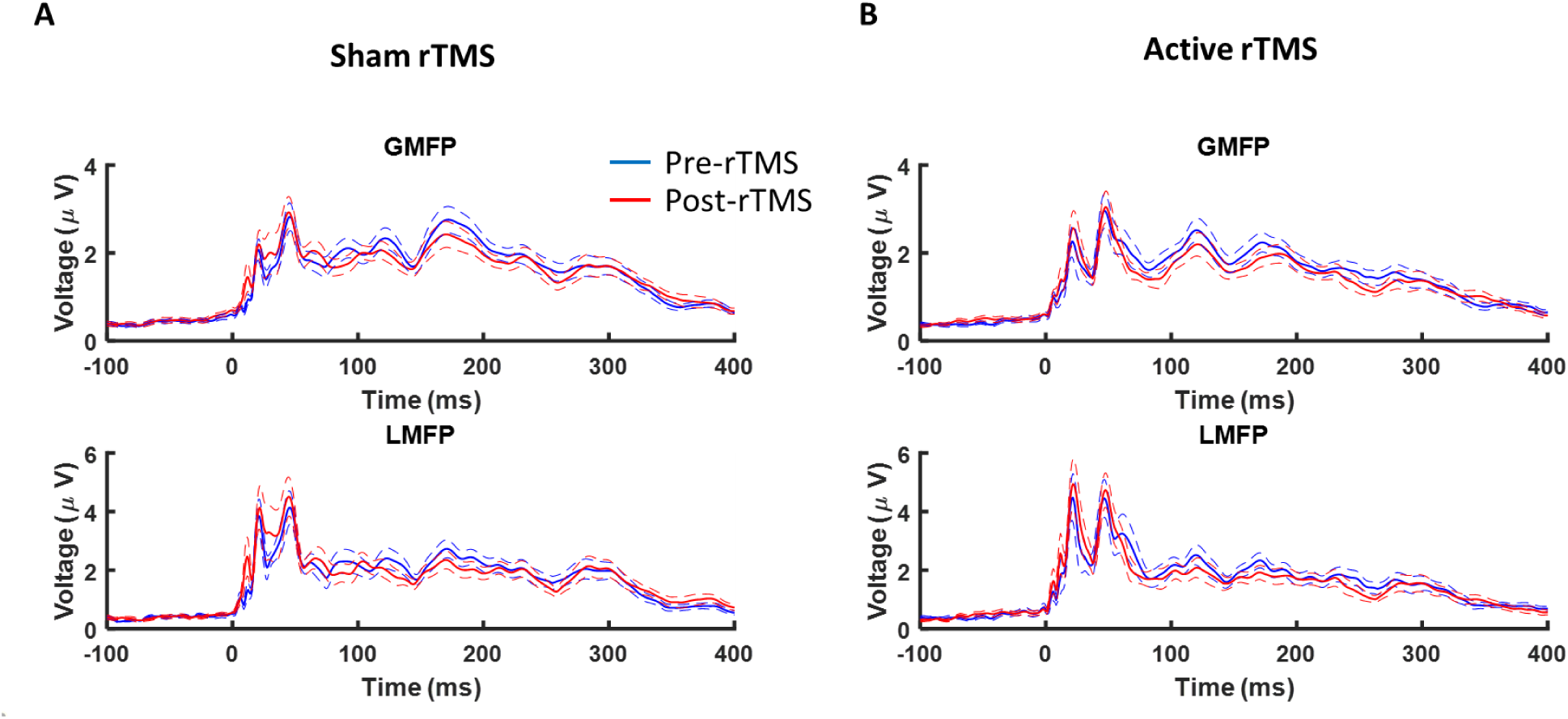
Global Mean Field Power (GMFP) and Local Mean Field Power (LMFP) from the dorsolateral prefrontal cortex (DLPFC) stimulation. Before repetitive transcranial magnetic stimulation (Pre- rTMS) is shown in blue, and immediately after repetitive transcranial magnetic stimulation (post- rTMS) is in red. Solid lines indicate the mean values, and dashed lines represent the standard deviation for each condition. **A)** Sham rTMS 10 Hz rTMS to M1; **B)** Active rTMS 10 Hz rTMS to M1.

**Supplementary figure 3.**
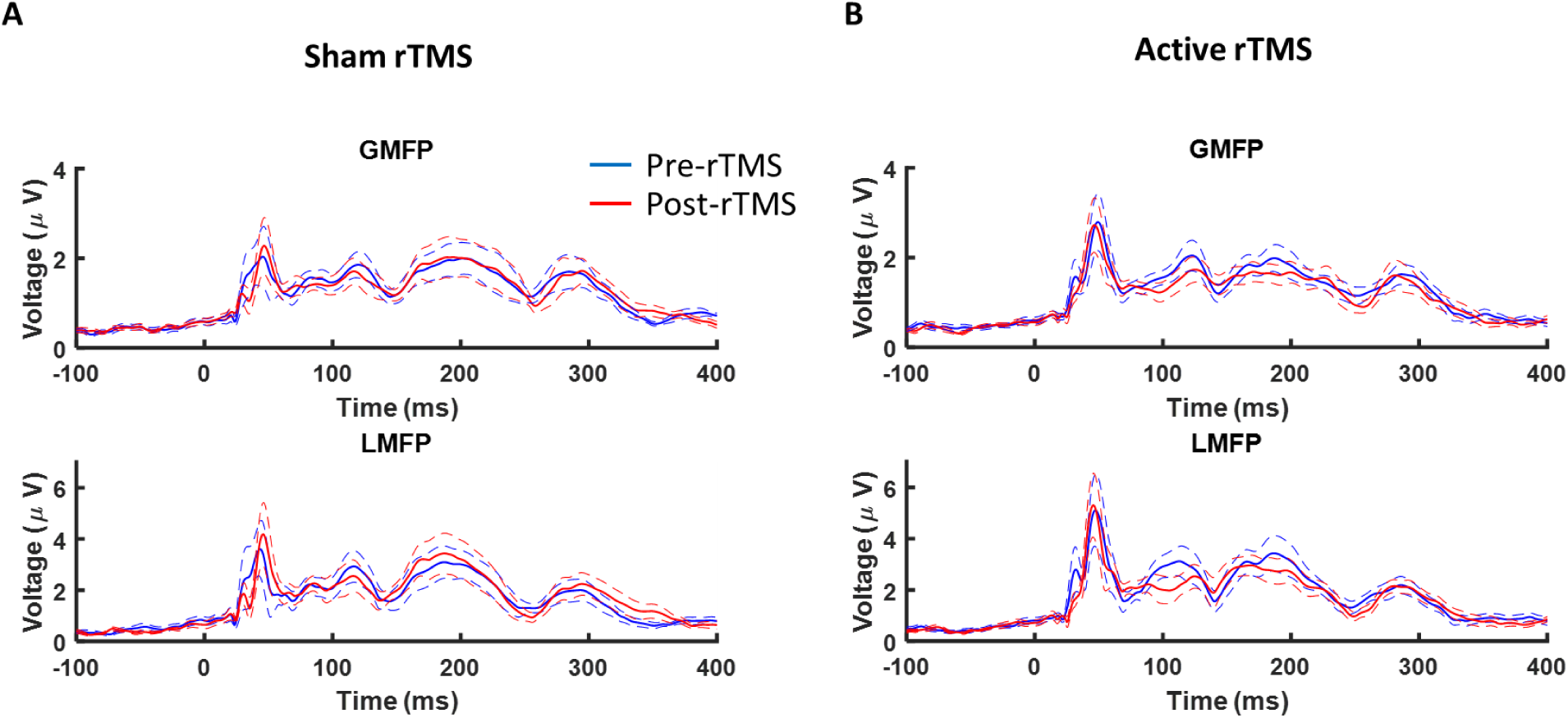
Global Mean Field Power (GMFP) and Local Mean Field Power (LMFP) from the anterior cingulate cortex (ACC) stimulation. Before repetitive transcranial magnetic stimulation (Pre-rTMS) is shown in blue, and immediately after repetitive transcranial magnetic stimulation (post-rTMS) is in red. Solid lines indicate the mean values, and dashed lines represent the standard deviation for each condition. **A)** Sham rTMS 10 Hz rTMS to M1; **B)** Active rTMS 10 Hz rTMS to M1.

**Supplementary figure 4.**
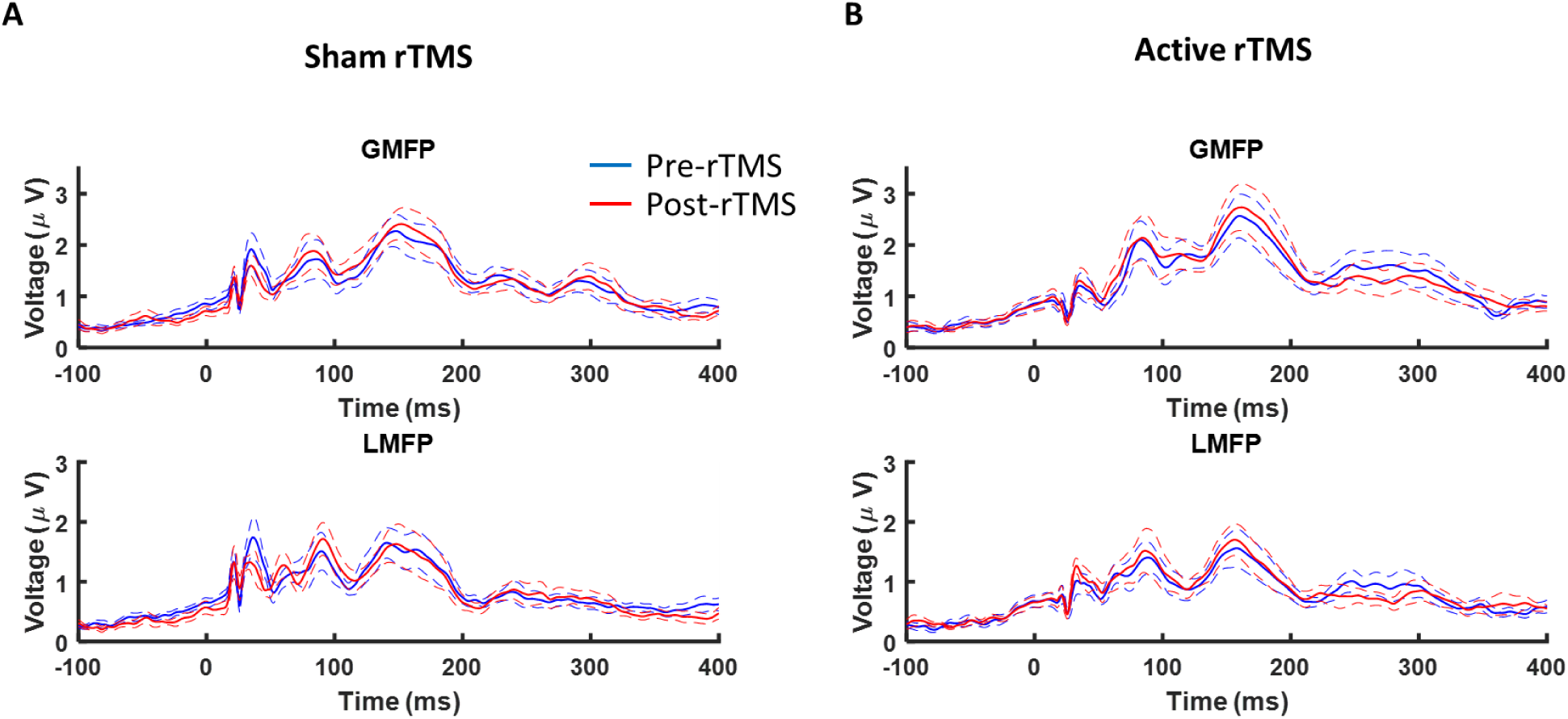
Global Mean Field Power (GMFP) and Local Mean Field Power (LMFP) from the posterosuperior insula cortex (PSI) stimulation. Before repetitive transcranial magnetic stimulation (Pre-rTMS) is shown in blue, and immediately after repetitive transcranial magnetic stimulation (post-rTMS) is in red. Solid lines indicate the mean values, and dashed lines represent the standard deviation for each condition. **A)** Sham rTMS 10 Hz rTMS to M1; **B)** Active rTMS 10 Hz rTMS to M1.

**Supplementary Table 1.** Mean ± standard deviation of Transcranial Magnetic Stimulation (TMS) power intensity used for evoking TMS-evoked potentials for each cortical spot and for sham and active repetitive transcranial magnetic stimulation (rTMS). For M1 and DLPFC, a figure-of-eight coil was set to the first dorsal interosseous muscle rTMS, and for ACC and PSI, a double-cone coil was set to the tibialis anterior muscle rTMS.

| Area | Condition | Maximum stimulator output |
| --- | --- | --- |
| M1 | Sham | 59.8%±6.9 |
| | Active | 60.1%±7.0 |
| DLPFC | Sham | 73.1%±8.6 |
| | Active | 73.5%±8.2 |
| ACC | Sham | 37.9%±6.1 |
| | Active | 38.1%±6.0 |
| PSI | Sham | 40.8%±3.3 |
| | Active | 40.3%±3.7 |

M1 = motor cortex; DLPFC = dorsolateral prefrontal cortex; ACC = anterior cingulate cortex; PSI = postero-superior insula cortex

**Supplementary Table 2.** Mean ± standard deviation of the average number of artifact-free epochs for each condition and for each cortical spot for sham and repetitive transcranial magnetic stimulation (rTMS).

| Area | Condition | Pre-rTMS | Post-rTMS |
| --- | --- | --- | --- |
| M1 | Sham | 148±16 | 149±16 |
| | Active | 151±16 | 151±16 |
| DLPFC | Sham | 151±21 | 150±18 |
| | Active | 153±19 | 152±15 |
| ACC | Sham | 141±15 | 138±7 |
| | Active | 138±18 | 137±15 |
| PSI | Sham | 157±6 | 155±10 |
| | Active | 159±8 | 158±7 |

M1 = motor cortex; DLPFC = dorsolateral prefrontal cortex; ACC = anterior cingulate cortex; PSI = postero-superior insula cortex

**Supplementary Table 3.** Mean ± standard deviation of the motor-evoked potentials before and after active and sham repetitive Transcranial Magnetic Stimulation (rTMS) to M1 at 120% and 140% of resting motor threshold.

| Variable | Condition | Before rTMS | After rTMS |
| --- | --- | --- | --- |
| **Motor-evoked potentials** |  |  |  |
| MEPs 120% (μV) | Sham rTMS | 460.3±393.3 | 596.3±548.8 |
| | Active rTMS | 616.2±489.4 | 657.8±647.6 |
| MEPs 140% (μV) | Sham rTMS | 963.3±609.8 | 1091.8±801.8 |
| | Active rTMS | 1150.2±800.5 | 1166.8±904.5 |

