## Supplementary material for "Repetitive transcranial magnetic stimulation to the motor cortex leads to a sequential increase in phase synchronization and power of TMS-evoked electroencephalographic recordings"

**Supplementary figure 1**

Global Mean Field Power (GMFP) and Local Mean Field Power (LMFP) from the primary motor cortex (M1) stimulation. Before repetitive transcranial magnetic stimulation (Pre-rTMS) is shown in blue, and immediately after repetitive transcranial magnetic stimulation (post-rTMS) is in red. Solid lines indicate the mean values, and dashed lines represent the standard deviation for each condition. **A)** Sham rTMS 10 Hz rTMS to M1; **B)** Active rTMS 10 Hz rTMS to M1.

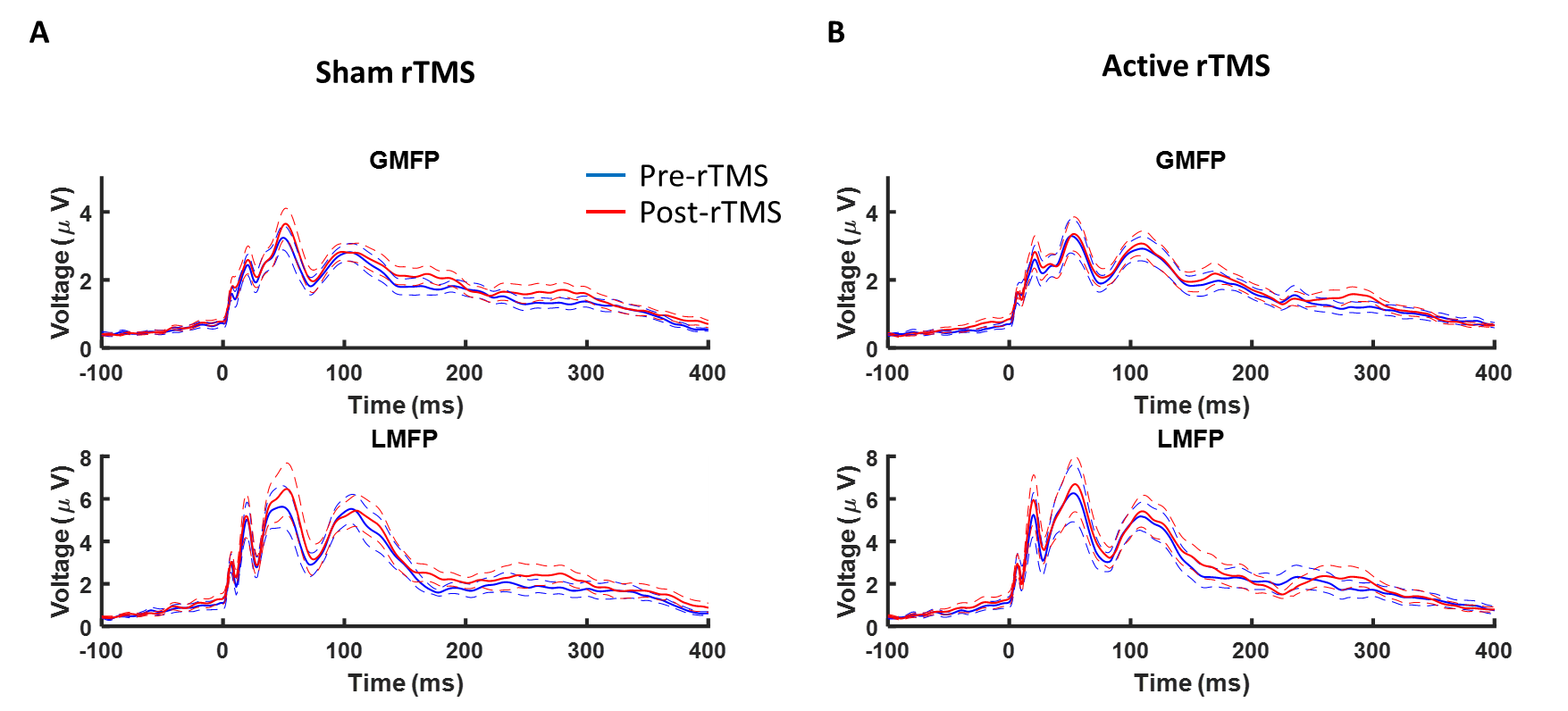

**Supplementary figure 2**

Global Mean Field Power (GMFP) and Local Mean Field Power (LMFP) from the dorsolateral prefrontal cortex (DLPFC) stimulation. Before repetitive transcranial magnetic stimulation (Pre-rTMS) is shown in blue, and immediately after repetitive transcranial magnetic stimulation (post-rTMS) is in red. Solid lines indicate the mean values, and dashed lines represent the standard deviation for each condition. **A)** Sham rTMS 10 Hz rTMS to M1; **B)** Active rTMS 10 Hz rTMS to M1.

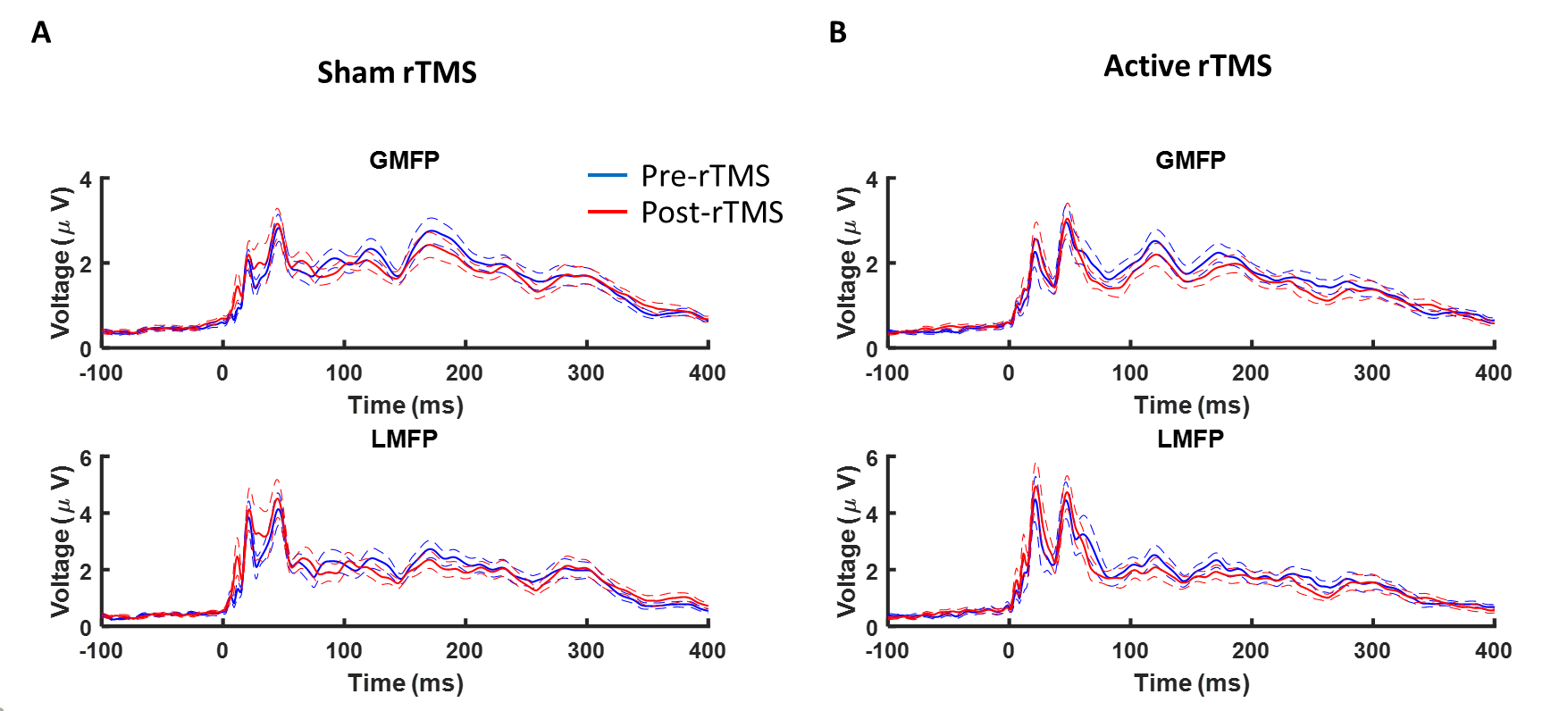

**Supplementary figure 3**

Global Mean Field Power (GMFP) and Local Mean Field Power (LMFP) from the anterior cingulate cortex (ACC) stimulation. Before repetitive transcranial magnetic stimulation (Pre-rTMS) is shown in blue, and immediately after repetitive transcranial magnetic stimulation (post-rTMS) is in red. Solid lines indicate the mean values, and dashed lines represent the standard deviation for each condition. **A)** Sham rTMS 10 Hz rTMS to M1; **B)** Active rTMS 10 Hz rTMS to M1.

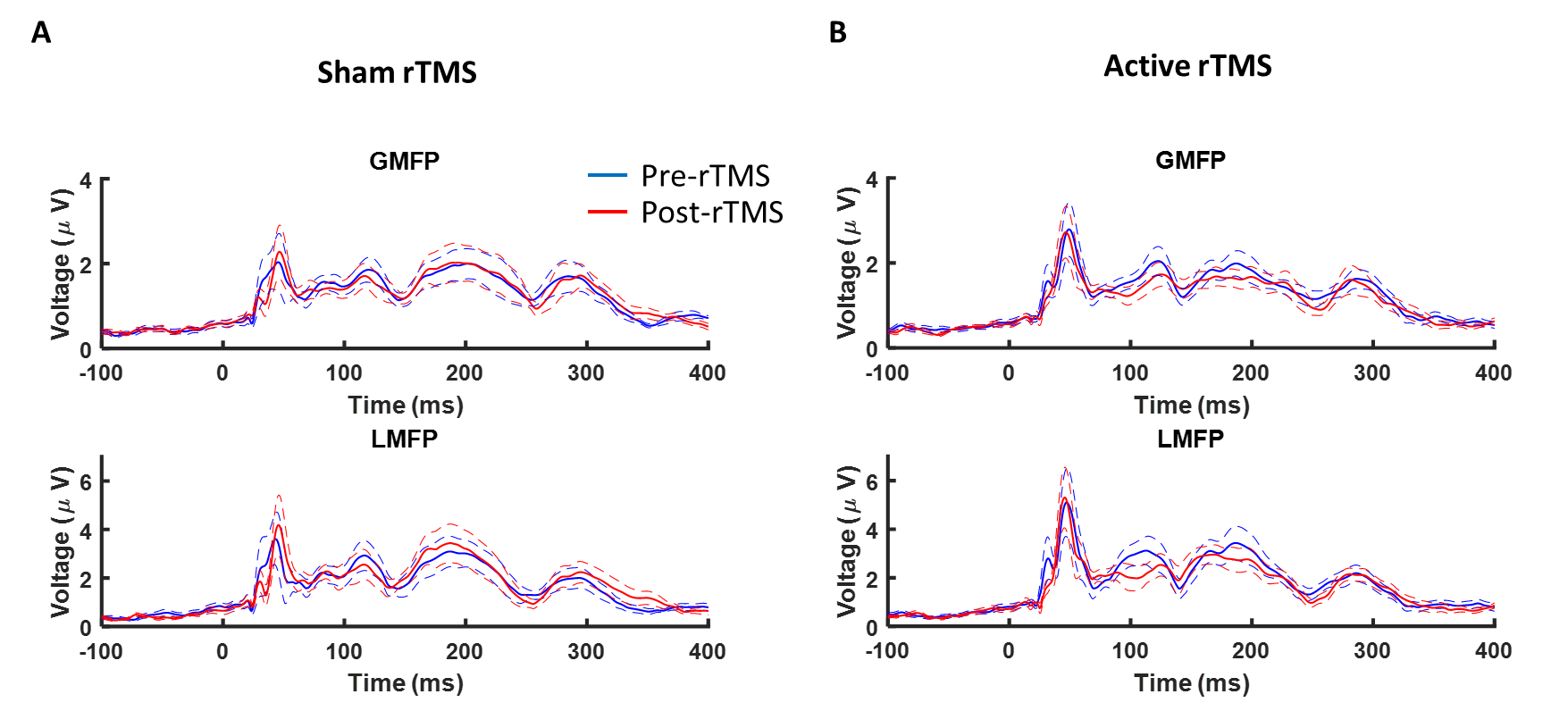

**Supplementary figure 4**

Global Mean Field Power (GMFP) and Local Mean Field Power (LMFP) from the posterosuperior insula cortex (PSI) stimulation. Before repetitive transcranial magnetic stimulation (Pre-rTMS) is shown in blue, and immediately after repetitive transcranial magnetic stimulation (post-rTMS) is in red. Solid lines indicate the mean values, and dashed lines represent the standard deviation for each condition. **A)** Sham rTMS 10 Hz rTMS to M1; **B)** Active rTMS 10 Hz rTMS to M1.

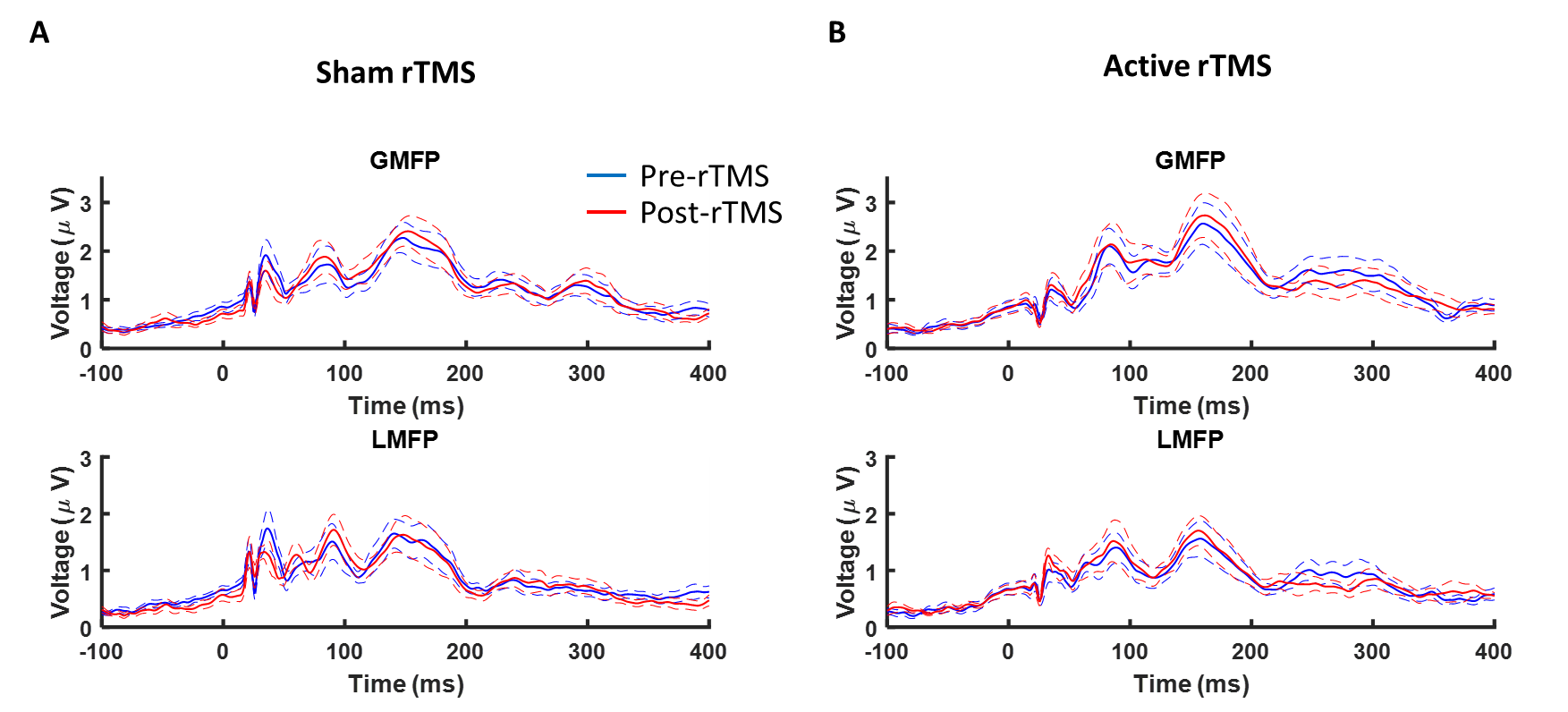

**Supplementary Table 1.** Mean ± standard deviation of Transcranial Magnetic Stimulation (TMS) power intensity used for evoking TMS-evoked potentials for each cortical spot and for sham and active repetitive transcranial magnetic stimulation (rTMS). For M1 and DLPFC, a figure-of-eight coil was set to the first dorsal interosseous muscle rTMS, and for ACC and PSI, a double-cone coil was set to the tibialis anterior muscle rTMS.

| **Area** | **Condition** | **Maximum stimulator output** |
| --- | --- | --- |
| M1 | Sham | 59.8%±6.9 |
|  | Active | 60.1%±7.0 |
| DLPFC | Sham | 73.1%±8.6 |
|  | Active | 73.5%±8.2 |
| ACC | Sham | 37.9%±6.1 |
|  | Active | 38.1%±6.0 |
| PSI | Sham | 40.8%±3.3 |
|  | Active | 40.3%±3.7 |

M1 = motor cortex; DLPFC = dorsolateral prefrontal cortex; ACC = anterior cingulate cortex; PSI = postero-superior insula cortex

**Supplementary Table 2.** Mean ± standard deviation of the average number of artifact-free epochs for each condition and for each cortical spot for sham and repetitive transcranial magnetic stimulation (rTMS).

| **Area** | **Condition** | **Pre-rTMS** | | **Post-rTMS** |
| --- | --- | --- | --- | --- |
| M1 | Sham | 148±16 | 149±16 | |
|  | Active | 151±16 | 151±16 | |
| DLPFC | Sham | 151±21 | 150±18 | |
|  | Active | 153±19 | 152±15 | |
| ACC | Sham | 141±15 | 138±7 | |
|  | Active | 138±18 | 137±15 | |
| PSI | Sham | 157±6 | 155±10 | |
|  | Active | 159±8 | 158±7 | |

M1 = motor cortex; DLPFC = dorsolateral prefrontal cortex; ACC = anterior cingulate cortex; PSI = postero-superior insula cortex

**Supplementary Table 3.** Mean ± standard deviation of the motor-evoked potentials before and after active and sham repetitive Transcranial Magnetic Stimulation (rTMS) to M1 at 120% and 140% of resting motor threshold.

| **Variable** | **Condition** | | **Before rTMS** | **After rTMS** |
| --- | --- | --- | --- | --- |
| **Motor-evoked potentials** | | | | |
| MEPs 120% (μV) | | Sham rTMS | 460.3±393.3 | 596.3±548.8 |
|  |  | Active rTMS | 616.2±489.4 | 657.8±647.6 |
| MEPs 140% (μV) | | Sham rTMS | 963.3±609.8 | 1091.8±801.8 |
|  |  | Active rTMS | 1150.2±800.5 | 1166.8±904.5 |
